# Unravelling the Links Between Chronotype, Body Mass Index, and Self-Regulatory Eating Behavior: Preliminary Insights from an Urban Kolkata Study

**DOI:** 10.1101/2025.04.02.25325081

**Authors:** Sayani Das, Sangita Mazumder, Kaninika Roy

## Abstract

**Background:** The rapid urbanization and changing lifestyles in India have led to an increase in obesity and metabolic disorders. Variations in chronotype can affect metabolic health and eating habits; however, there is a scarcity of research examining their relationship with body mass index (BMI) and self-regulatory eating behavior (SREB) within Indian populations. This study aims to investigate these associations in a cohort from urban Kolkata.

**Methods:** A cross-sectional study was performed involving 156 adults aged 18 to 60 years in West Bengal. The reduced Morningness-Eveningness Questionnaire (rMEQ) was used to determine chronotype, whereas the Self-Regulation of Eating Behavior Questionnaire (SREBQ) was applied to assess self-regulation of eating behavior (SREB). Body Mass Index (BMI) was categorized into underweight, normal weight, overweight, and obesity. To investigate the associations between chronotype, BMI, and SREB, chi-square tests and logistic regression analyses were performed.

**Results:** The distribution of chronotypes was as follows: morning (31%), intermediate (45%), and evening (24%). No significant correlation was identified between chronotype and BMI (p = 0.34). Evening chronotypes displayed lower SREB compared to morning and intermediate types; however, this difference was not statistically significant (p = 0.17). Logistic regression analysis indicated that individuals with intermediate chronotypes had significantly greater odds of exhibiting self-regulation compared to those with evening chronotypes (OR: 7.77, p = 0.005). Participants with postgraduate education showed improved SREB (p = 0.037).

**Conclusion:** Individuals with intermediate chronotypes demonstrate superior self-regulation in eating behaviors compared to those with evening chronotypes, underscoring the importance of circadian alignment in dietary practices. No direct association was established between chronotype and BMI, indicating a need for further longitudinal research to inform targeted dietary interventions.

## Introduction

Urbanization and rapid lifestyle changes in India have led to a surge in obesity and related metabolic disorders. Concurrently, disrupted sleep patterns and altered circadian rhythms have become prevalent, raising concerns about their potential impact on weight management. Understanding how chronotype, as a marker of circadian preference, influences BMI and eating behaviors in this population could provide valuable insights into the development of targeted interventions for obesity prevention and management.

The circadian rhythm refers to the series of biological and metabolic processes that occur within a 24-hour period (Czeisler et al. 1999). The suprachiasmatic nucleus (SCN) in the hypothalamus plays a pivotal role in regulating this rhythm. Individuals can be categorized into three primary chronotype: Morning-types (M-types), Evening-types (E-types), and neither/Intermediate-types (N-types) (Horne and Östberg 1976). Morning chronotypes typically retire early in the evening and rise early in the morning, showcasing peak performance during the early hours. Conversely, evening chronotypes tend to stay up late, struggle with early wakeups, and exhibit higher performance later in the day. Several factors, including genetic makeup, age, ethnicity, and gender, influence an individual’s chronotype (Roenneberg et al. 2007).

Variations in chronotype can have significant health implications. Studies indicate that chronotype is linked to an increased risk of various illnesses, including metabolic syndrome, Type 2 Diabetes Mellitus (DM), cardiovascular diseases (CVD), and depression, as well as its impact on obesity (Almoosawi et al. 2019; Aoun et al. 2019; Hawley et al. 2020; Baron et al. 2011; Merikanto et al. 2014). Evening chronotypes, characterized by a delayed circadian phase, are associated with impaired glycaemic control, metabolic dysfunction, a higher risk of cardiovascular diseases, obesity, and psychiatric disorders (Reutrakul and Knutson 2015; Lucassen et al. 2013; Partonen 2015). Consequently, individuals with evening preferences are at higher risk of morbidity and mortality compared to those with morning preferences (Knutson and von Schantz 2018). This increased risk is frequently linked to chronic circadian misalignment, where an individual’s internal clock is not synchronized with external work and social demands. Evidence suggests that evening chronotypes tend to have a higher BMI, poorer dietary habits, and lower levels of physical activity (Lucassen et al. 2013).

Recently, differences in susceptibility to environments that promote obesity have become more apparent (Blundell et al. 2020; Wells and Siervo 2011). It is proposed that a person’s capacity for self-regulation and their ability to resist food temptations may significantly influence this variation., potentially leading to healthier weight management and dietary habits. Research indicates that effective self-regulation of eating behaviors can bridge the gap between one’s intentions and actual behaviors, thus supporting the achievement of dietary goals (Kliemann 2016).

Various studies have extensively explored the connections between chronotype, nutrients, and dietary habits. Research indicates that individuals with evening chronotypes are more inclined to skip meals, consume lower amounts of fruits and vegetables, indulge in high sugary drinks, have high alcohol intake, and struggle with controlling portion sizes (Kanerva et al. 2012). These findings raise the question of whether evening chronotypes exhibit inadequate self-regulation of eating behavior, resulting in the adoption of unhealthy dietary patterns. Self-regulation involves the ability to manage one’s behaviors, thoughts, emotions, and surroundings to support the achievement of specific goals (Kliemann 2016).

The evidence presented lays the groundwork for suggesting that individuals with evening chronotypes may struggle to regulate their eating habits. In the bustling metropolis of Kolkata, balancing environmental factors such as work schedules and social commitments often complicates the maintenance of a healthy diet. Nonetheless, developing self-regulation skills could potentially lead to better dietary practices and healthy body weight. To explore this theory, it is essential to examine the connections between various chronotypes and the ability to resist tempting foods in relation to body weight. As far as we are aware, very few research has delved into the link between chronotype and self-regulatory eating behaviour in Indian population, particularly in Kolkata. Hence, the primary objective of this research is to evaluate the correlations between chronotype, BMI and the self-regulation of eating behavior among adults of Kolkata.

## Methods

### Study design and participants

This cross-sectional study was conducted in different part of West Bengal with the participation of 156 adults between September 2023 to May 2024. Exclusion criteria included individuals under 18 years of age and those over 60 years, as well as pregnant or breastfeeding participants, individuals with chronic illnesses, those taking any medications, and those diagnosed with psychiatric disorders. The questionnaire was made available in English and disseminated through a Google Form link via WhatsApp and email. The rMEQ questionnaires and SREB questionnaires and demographic questionnaire are included. Both close ended questionnaire and open-ended questionnaire are included.

### Power Analysis

To ensure that our study was adequately powered to detect significant associations, we conducted a power analysis. Using a medium effect size (0.5), a significance level (alpha) of 0.05, and a desired power of 0.8, the analysis indicated that a sample size of approximately 64 participants was required. Our study included 156 participants, which is well above the required sample size. This suggests that our study is sufficiently powered to identify significant relationships between chronotype, BMI, and self-regulatory eating behavior.

### rMorningness-Eveningness Questionnaire

The Morningness–Eveningness Questionnaire (MEQ) is widely recognized as the standard tool for assessing chronotype (Horne and Östberg 1976). Developed by Horne and Östberg in 1976, the MEQ has undergone rigorous validation and is extensively utilized in research (Horne and Östberg 1976). To enhance participant compliance, a shorter 5-item version known as the reduced Morningness–Eveningness Questionnaire (rMEQ) was employed in the current study. The rMEQ has demonstrated reliability, good psychometric properties, and convergent validity (Adan and Almirall H 1991). Comprising five Likert-type questions, the reduced questionnaire generates scores ranging from 4 to 25. Questions 1 to 3 inquire about participants’ preferred wake-up time, subjective alertness levels during the day and night, while Question 4 assesses the time of day when they feel most productive. Question 5 directly addresses their morningness or eveningness preferences. The overall score, which spans from 4 (indicating extreme eveningness) to 25 (indicating extreme morningness), categorizes individuals into five distinct chronotype classifications: ‘definitely morning’ type, ‘moderately morning’ type, ‘neither (intermediate)’ type, ‘moderately evening’ type, and ‘definitely evening’ type (Adan and Almirall H 1991). Consistent with earlier studies, the classifications were simplified into morning type, neither (intermediate) type, and evening type to avoid the issue of having categories with insufficient sample sizes, as highlighted by various research findings (Adan et al. 2010; Culnan et al. 2013; Sun et al. 2020)

### Self-Regulation of Eating Behavior Questionnaire (SREBQ)

The Self-Regulation of Eating Behavior Questionnaire (SREBQ) is employed to assess an individual’s capacity for self-regulating their eating habits. This questionnaire has undergone validation and has demonstrated strong reliability in achieving its intended objectives (Kliemann 2016). The assessment comprises five items with response options that range from 1 (never) to 5 (always). It begins with a list of 13 frequently desired foods, followed by the inquiry: “Do you find any of the following foods appealing (meaning, do you wish to consume more of them than you believe is appropriate)?” Participants can select all the foods they find appealing. The next two questions ask whether they plan to manage their intake of these appealing foods and if they intend to follow a healthy diet. These questions aim to filter out participants who lack healthy dietary intentions from the SREBQ. As a result, only those with healthy dietary intentions are permitted to complete the SREBQ, and their ability to self-regulate in maintaining these intentions is subsequently assessed. The mean score thresholds are as follows: a score of less than 2.8 indicates low self-regulation, a score between 2.8 and 3.6 indicates moderate self-regulation, and a score above 3.6 indicates high self-regulation of eating behavior (Kliemann 2016).

### Self-structured question for Demographic Data

The questionnaire incorporated socio-economic factors and personal inquiries. It included questions regarding individuals’ personal information such as name, age, gender (male/female), socio-economic factors like educational attainment (e.g., never attended school, primary or intermediate school, high school, diploma, graduate or master’s degree), as well as employment status (employee or non-employee) and shift work. All participants provided their consent and agreed to disclose their personal information.

### Anthropometric Measurement

Participant’s weight and height were reported by the participants. BMI was calculated by formula weight (kg) divided by height (m)^2^. It is used as a measure of fatness in adult. The BMI of participants were derived into six categories. Below 18.5 kg/m^2^ (Underweight), 18.5-22.9 kg/m^2^ (Normal weight), 23.0-24.9 kg/m^2^ (Overweight), 25-29.9 kg/m^2^ (obese I), and above 30 kg/m^2^ (obese II) (WHO Expert Consultation 2004).

### Statistical analysis

Data was utilized by using the statistical package for the social sciences (SPSS). Descriptive statistics like Frequency and percentages were used to demonstrate the demographical data, that is age of students, gender, education of participants, employment and shiftwork of participants. BMI are also described by using percentage. Mean and standard deviation were used to present numerical variable. Chi-square test was also used to calculated analysis about association between chronotype and BMI, shift work with BMI, and chronotype with self - regulatory eating behaviour as well. This association reveals the hypothetical situation about this factor among 18-60years of age. Eligibility for the SREB required participants to express a commitment to maintaining a healthy diet. Consequently, those who do not intend to follow a healthy dietary regimen should be categorized separately. The significance level was established at 0.05.

## Results

### Socio-Demographic characteristics

A total of 156 individuals completed the survey questionnaire. The socio-demographic profile of the participants is presented in Table 1. Out of the total participants, 112 (71.8%) were aged between 18 and 29 years, 23 (14.7%) were aged between 30 and 40 years, 21 (13.4%) were aged between 41 and 60 years. Both male and female participants were included in the study, with 96 (61.5%) females and 60 (38.5%) males. The majority of the participants were female, accounting for 61.5% of the total. Each participant was involved in various levels of education, such as undergraduate studies, pursuing a Master of Science degree, etc. The distribution of educational levels among the participants was as follows: 53.2% were graduates, 33.3% were postgraduates, and 13.5% held a HS degree. In terms of occupation, 23.1% of the participants were students, 34.6% were employed, and 42.3% were unemployed. Regarding Body Mass Index (BMI), the majority of participants, accounting for 32.7%, fell into the obese category. 13.5% were classified as underweight, 29.5% were in the normal and 24.4% were in overweight categories.

**Table 1:** Demographic profile of participants (n=156)

| Demographical Characteristics | Frequency | Percent (%) |
| --- | --- | --- |
| **AGE** |  |  |
| 18y-29y | 112 | 71.8% |
| 30y-40y | 23 | 14.7% |
| 41-60 y | 21 | 13.4% |
| **GENDER** |  |  |
| M | 60 | 38.5% |
| F | 96 | 61.5% |
| **EDUCATION** |  |  |
| HS School | 21 | 13.5% |
| Graduation | 83 | 53.2% |
| Post-graduation and above | 52 | 33.3% |
| **EMPLOYEMENT** |  |  |
| Employed | 54 | 34.6% |
| Unemployed | 66 | 42.3% |
| Student | 36 | 23.1% |
| **BMI** |  |  |
| Normal | 46 | 29.5% |
| Underweight | 21 | 13.5% |
| Overweight | 38 | 24.4% |
| Obese | 51 | 32.7% |
| **Having healthy dietary intension** | 156 | 100% |

**BMI = body mass index, n = number.**

### Chronotype and associated factor

Table 2 displays the distribution of chronotype among the participants. Based on rMEQ questionnaire, 31% participants were classified as morning chronotype, 24% as the evening chronotype, and the majority, 45%, fell into the intermediate or neither type category, meaning they are neither morning nor evening chronotype. (n= 48; n= 38; n= 70 respectively).

**Table 2:** Factors related to chronotype (n=156)

| Factor | Chronotype |  |  | p value |
| --- | --- | --- | --- | --- |
|  | Morning | Intermediate | Evening |  |
| BMI |  |  |  |  |
| Underweight | 7 (14.6%) | 5 (7.1%) | 9 (23.7%) | 0.34 |
| Normal | 13 (27.1%) | 21 (30%) | 12 (31.6%) |  |
| Overweight | 12 (25%) | 18 (25.7%) | 8 (21.1%) |  |
| Obese | 16 (33.3%) | 26 (37.1%) | 9 (23.7%) |  |
| Age |  |  |  |  |
| 18-29 years | 34 (70.8%) | 49 (70%) | 29 (76.3%) | 0.023* |
| 30-40 years | 3 (6.3%) | 16 (22.9%) | 4 (10.5%) |  |
| 41-60 years | 11 (22.9%) | 5 (7.1%) | 5 (7.1%) |  |
| Self-regulation of eating bahavior |  |  |  |  |
| Low | 9 (18.8%) | 17 (24.3%) | 10 (26.3%) | 0.17 |
| Medium | 31 (64.6%) | 35 (50%) | 25 (65.8%) |  |
| High | 8 (16.7%) | 18 (25.7%) | 3 (7.9%) |  |
| Sex |  |  |  |  |
| Male | 23 (47.9%) | 25 (35.7%) | 12 (31.6%) | 0.25 |
| Female | 25 (52.1%) | 45 (64.3%) | 26 (68.4%) |  |
| Education |  |  |  |  |
| Higher secondary | 6 (12.5%) | 12 (17.1%) | 3 (7.9%) | 0.23 |
| Graduation | 29 (60.4%) | 37 (52.9%) | 17 (44.7%) |  |
| Post-graduation | 13 (27.1%) | 21 (30%) | 18 (47.4%) |  |
| Employment |  |  |  |  |
| Students | 6 (12.5%) | 15 (21.4%) | 15 (39.5%) | 0.036* |
| Employed | 17 (35.4%) | 24 (34.3%) | 13 (34.2%) |  |
| Unemployed | 25 (52.1%) | 31 (44.3%) | 10 (26.3%) |  |

ANOVA and Chi-square tests were employed.
The thresholds for self-regulation of eating behavior are categorized as follows: low is defined <2.8, medium ranges from 2.8 to 3.6, and high is >3.6.

Table 2 further investigates the connection between Body Mass Index (BMI), self-regulatory eating behavior (SREB), and chronotype. A majority of participants displayed medium self-regulation (n=91; 58.3%). Those with evening chronotypes showed a greater inclination towards low self-regulation (26.3%) in comparison to morning (18.8%) and intermediate types (24.3%), although this difference did not reach statistical significance (p=0.17). There were no significant variations in BMI among the different chronotype categories. Moreover, factors such as gender and education level did not correlate with chronotype, while age and employment status were found to be associated with it.

### Self-regulation of Eating Behavior Questionnaire (SREBQ)

The questionnaire revealed that all participants had healthy dietary intentions, resulting in valid SREBQ scores. The participants’ SREBQ scores indicated that 23.1% (n = 36) had low self-regulation of eating behavior, 58.3% (n = 91) had moderate self-regulation, and 18.5% (n = 29) had high self-regulation. The participants’ responses to the SREBQ are detailed in Table 3.

**Table 3:** Responses from participants regarding the Self-Regulatory Eating Behavior Questionnaires (SREBQ) (n=156)

| **Question** | **n** | **Percentage (%)** |
| --- | --- | --- |
| **I give up too easily on my eating intentions** |  |  |
| Always | 3 | 1.9 |
| Often | 20 | 12.8 |
| Sometimes | 77 | 49.3 |
| Rarely | 44 | 28.2 |
| Never | 12 | 7.7 |
| **I'm good at resisting tempting food** |  |  |
| Always | 17 | 10.9 |
| Often | 25 | 16.02 |
| Sometimes | 68 | 43.6 |
| Rarely | 35 | 22.4 |
| Never | 11 | 7.05 |
| **I easily get distracted from the way I intend to eat** |  |  |
| Always | 6 | 3.8 |
| Often | 21 | 13.5 |
| Sometimes | 52 | 33.3 |
| Rarely | 48 | 30.8 |
| Never | 29 | 18.6 |
| **If I am not eating in the way, I intend to I make changes** |  |  |
| Always | 7 | 4.5 |
| Often | 15 | 9.6 |
| Sometimes | 60 | 38.5 |
| Rarely | 45 | 28.9 |
| Never | 29 | 18.6 |
| **I find it hard to remember what I have eaten** |  |  |
| Always | 6 | 3.8 |
| Often | 22 | 14.1 |
| Sometimes | 43 | 27.6 |
| Rarely | 32 | 20.5 |
| Never | 53 | 34 |

Figure 1 illustrates that 12.8% (n = 20) of participants reported not finding any food tempting. The most tempting foods were chocolate (n = 78; 50%), pizza (n = 57; 36.5%), sweets (n = 50; 32%), and fried foods (n = 48; 30.8%).

**Figure 1.**
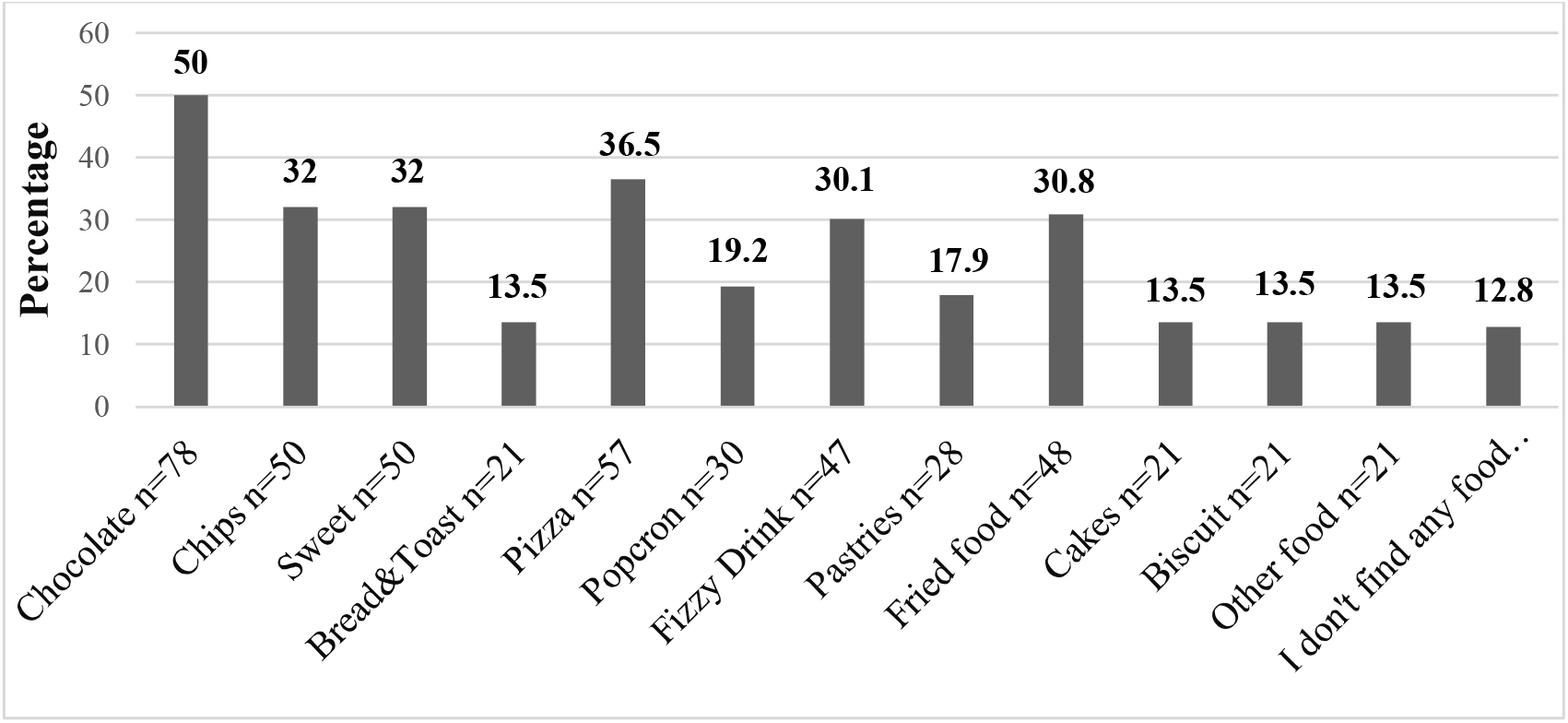
Responses from the Participants to the inquiry “do you find any of the following food tempting (That is, do you want to eat more of them than you think you should)?”

### Chronotype, Gender, Employment, BMI and SREB

Table 4 illustrates the relationship between food items that participants find tempting and their chronotype and gender. The results indicate a notable correlation between the temptation of soft drinks and chronotype (p = 0.03). While not statistically significant, individuals with an evening chronotype showed a greater tendency to find foods such as chocolate, bread/toast, pastries, fried foods, and cake appealing and hard to resist compared to those with a morning or intermediate chronotype. Conversely, participants with a morning chronotype exhibited the least vulnerability to these temptations.

**Table 4:** Association between the types of foods that participants find appealing, as influenced by their chronotype and gender.

| Food participants find to be tempting | Chronotype |  |  | p value |
| --- | --- | --- | --- | --- |
|  | Morning<br>n (% from morning type) | Intermediate<br>n (% from intermediate type) | Evening<br>n (% from evening type) |  |
| **Chocolate** | 19 (39.6%) | 36 (51.4%) | 23 (60.5%) | 0.15 |
| **Chips** | 10 (20.8%) | 26 (37.1%) | 14 (36.8%) | 0.14 |
| **Sweets** | 17 (35.4%) | 24 (34.3%) | 9 (23.7%) | 0.44 |
| **Bread/ toast** | 6 (12.5%) | 7 (10%) | 8 (21.1%) | 0.27 |
| **Pizza** | 16 (33.3%) | 27 (38.6%) | 14 (36.8%) | 0.84 |
| **Popcorn** | 8 (16.7%) | 16 (22.9%) | 6 (15.8%) | 0.58 |
| **Frizzy drink** | 11 (22.9%) | 18 (25.7%) | 18 (47.3%) | **0.03\*** |
| **Pastries** | 8 (16.7%) | 12 (17.1%) | 8 (21.1%) | 0.85 |
| **Fried food** | 13 (27.1%) | 19 (27.1%) | 16 (42.1%) | 0.22 |
| **Cake** | 5 (10.4%) | 8 (11.4%) | 8 (21.1%) | 0.29 |
| **Biscuit** | 7 (14.6%) | 9 (12.9%) | 5 (13.2%) | 0.96 |

| Food participants find to be tempting | Sex |  | p value |
| --- | --- | --- | --- |
|  | Male | Female |  |
| **Chocolate** | 22 (36.7%) | 56 (62.2%) | **0.008\*** |
| **Chips** | 15 (25%) | 35 (38.8%) | 0.136 |
| **Sweets** | 18 (30%) | 32 (35.5%) | 0.66 |
| **Bread/ toast** | 4 (6.7%) | 17 (18.8%) | **0.049\*** |
| **Pizza** | 20 (33.3%) | 37 (41.1%) | 0.51 |
| **Popcorn** | 7 (11.7%) | 23 (25.5%) | 0.059 |
| **Frizzy drink** | 16 (26.7%) | 31 (34.4%) | 0.456 |
| **Pastries** | 5 (8.3%) | 23 (25.5%) | 0.13 |
| **Fried food** | 13 (21.7%) | 35 (38.8%) | 0.051 |
| **Cake** | 13 (21.7%) | 8 (8.8%) | **0.018\*** |
| **Biscuit** | 7 (11.7%) | 14 (15.5%) | 0.60 |

Chi-square test used.
\*Significant at level 0.05

**Table 5:** Factors that predict high self-regulation of eating behavior.

| **Variables** | **Categories** | **S.E.** | **P value** | **Odds ratio** | **95% C.I.** |
| --- | --- | --- | --- | --- | --- |
| **Age** | 18-29 | 0.698 | 0.750 | 0.800 | (0.204,3.143) |
|  | 30-40 | 1.237 | 0.086 | 0.120 | (0.011,1.352) |
|  | 41-60 | Reference |  |  |  |
| **Gender** | Male | 0.531 | 0.230 | 1.891 | (0.668,5.352) |
|  | Female |  |  | Reference |  |
| **BMI** | Underweight | 0.730 | 0.444 | 1.749 | (0.418,7.316) |
|  | Normal | 0.600 | 0.564 | 1.414 | (0.436,4.587) |
|  | Overweight | 0.654 | 0.782 | 0.835 | (0.232,3.005) |
|  | Obese | Reference |  |  |  |
| **Education** | Higher secondary | 1.182 | 0.037* | 0.086 | (0.008,0.867) |
|  | Graduation | 0.512 | 0.579 | 0.753 | (0.276, 2.052) |
|  | Post-graduation or above | Reference |  |  |  |
| **Employment** | Students | 0.584 | 0.415 | 1.609 | (0.512,5.053) |
|  | Employed | 0.587 | 0.738 | 0.822 | (0.260,2.594) |
|  | Unemployed |  |  | Reference |  |
| **Chronotype** | Morning | 0.774 | 0.178 | 2.837 | (0.623,12.926) |
|  | Intermediate | 0.736 | 0.005* | 7.7743 | (1.829,32.779) |
|  | Evening | Reference |  |  |  |

Multivariate Logistic Regression analyses were employed.
Self-regulation of eating behavior (SREB) is categorized based on scores, with low SREB defined as a score of $\leq 3.6$ and high SREB defined as a score $> 3.6$ .
\*Significant at level 0.05.

Moreover, Table 4 also analyzes the connection between gender and the perception of food temptations. Women were significantly more inclined than men to view chocolate (p = 0.008) and bread/toast (p = 0.049) as tempting. On the other hand, men rated cake as significantly more tempting than women did (p = 0.018).

Table S1 examines the relationship between BMI categories and the perception of tempting foods. With the exception of bread/toast (p = 0.049), no significant correlations were identified between BMI and the foods deemed tempting by participants.

Additionally, Table S1 investigates the link between employment status and food temptations. The analysis revealed no significant relationship between employment status and participants’ preferences for these foods.

### Factors that predict high self-regulation of eating behavior

A multivariate logistic regression analysis was performed to identify the factors predicting high self-regulation of eating behavior. Intermediate chronotypes have significantly higher odds of the self-regulation of eating behavior compared to evening types, which suggests that individuals with intermediate chronotypes might have different behavioural or physiological patterns influencing the outcome. Individuals with higher secondary education have significantly lower odds of high self-regulation in eating behavior compared to those with post-graduation or higher education (reference). Age, Gender, BMI, and Employment did not show significant associations with the SREB, indicating that they may not be strong predictors in this context.

## Discussion

This research constitutes a fundamental examination of the correlation between chronotype and self-regulatory eating behavior (SREB) in people residing in Kolkata. Our findings demonstrate that individuals with an intermediate chronotype display superior self-regulation in dietary habits compared to those with an evening preference. This indicates that individuals with intermediate chronotypes may encounter reduced circadian misalignment, facilitating enhanced self-regulation. Their adaptable alignment with societal patterns and everyday requirements may facilitate the preservation of healthy eating habits.

Our findings notably differ with research in Saudi Arabia, which revealed that morning types had much greater self-regulation in eating behaviors—85% more than evening types (Al-Hazmi and Noorwali 2023). Various variables may explain this disparity (de Ridder 2012). The limited sample size of our study may restrict the generalizability of the findings, necessitating more research with bigger cohorts. Furthermore, our study utilized a cross-sectional strategy, which collects data at a singular moment, hence constraining causal conclusions among chronotype, BMI, and SREB. Future longitudinal research may yield more profound insights into these interactions. Furthermore, genetic and ethnic variables undoubtedly affect chronotype (Roenneberg et al. 2007), and the scarcity of comprehensive study on Indian communities hinders direct comparisons with other demographic groups.

In addition to chronotype, our research showed schooling as a key predictor of elevated SREB. This indicates that elements outside circadian cycles, including cognitive awareness, lifestyle decisions, and socio-environmental factors, may significantly influence self-regulated eating patterns. This corresponds with developing viewpoints that self-regulation is not exclusively governed by chronotype but is influenced by a wider array of psychological and environmental factors (de Ridder 2012).

Our findings indicate that evening chronotypes are more inclined towards unhealthy meals, especially soft drinks, while morning chronotypes display reduced susceptibility to these temptations. This aligns with previous research (Kanerva 2012) that correlates eveningness with increased fat consumption and suboptimal dietary practices. A credible reason is the timing of food intake—individuals with an evening chronotype tend to postpone meals, which has been associated with a higher consumption of high-fat foods and decreased nutritional quality (Mazri et al. 2019; Lopez-Minguez et al. 2019). Epidemiological data indicate that early breakfast consumption correlates with increased fiber intake, while late-night eating is associated with elevated saturated fat consumption (Khare and Inman 2006; Roßbach et al. 2018).

Nevertheless, our study revealed no significant evidence connecting chronotype and BMI. This corresponds with prior studies demonstrating no association between chronotype and BMI. For example, multiple cross-sectional investigations, including a comprehensive Finnish study with 4421 individuals (Maukonen et al. 2016) and a Norwegian population-based study comprising 6413 participants (Johnsen et al. 2013), revealed no association between BMI and chronotype. An analysis of 36 studies indicates that whereas evening chronotypes frequently exhibit detrimental food habits, both chronotypes generally have comparable total calorie consumption (Mazri et al. 2019). In contrast, a subsequent study including 54 participants demonstrated that evening chronotypes exhibited a notable increase in BMI after 8 weeks, despite no correlation being seen at baseline (Culnan et al. 2013). These findings highlight the necessity for more research to elucidate the long-term effects of chronotype on weight regulation.

Furthermore, our research revealed that gender disparities influence food desires. Women indicated a broader array of meals as enticing, such as chips, ice cream, popcorn, and pastries, whereas men had a stronger inclination towards cake. This is consistent with current data indicating that emotional eating exhibits a stronger correlation with intuitive eating in women compared to males (Smith et al. 2020). One potential explanation is that women may encounter heightened appetites for chocolate and sweets, especially during the premenstrual phase (De Souza et al. 2018; Gorczyca et al. 2016; Hintze et al. 2017). Furthermore, studies indicate that exercise exerts a more pronounced appetite-suppressing effect in men compared to women (Drenowatz et al. 2017), potentially influencing disparities in food temptation and dietary self-regulation.

These findings offer significant insights into the interplay between chronotype, SREB, and dietary choices, while also underscoring the intricacy of regulating eating behaviors. Multiple elements, including circadian rhythms, psychological influences, lifestyle choices, and environmental conditions, combine to shape an individual’s capacity for dietary self-regulation (Engin 2017; Lowe et al. 2019). Comprehending these connections can facilitate the development of more efficacious dietary interventions and tailored nutrition programs.

Future research should investigate the interaction between chronotype, psychological characteristics, and environmental impacts to further the knowledge of eating behaviors. Although our sample size was adequate for the power analysis, longitudinal studies would be especially beneficial in clarifying the impact of chronotype on self-regulation over time and its interaction with lifestyle and behavioral patterns.

In conclusion, although our study could not establish a clear correlation between chronotype and self-regulatory eating behavior, the identification of intermediate chronotype as a predictor of enhanced self-regulation offers significant insights. These findings underscore the necessity for a more sophisticated approach to comprehending chronotype and its interplay with lifestyle factors. Understanding the significance of flexible and adaptable circadian rhythms in self-regulation may enhance dietary management and overall health promotion techniques.

## Data Availability

All data produced in the present work are contained in the manuscript

## Acknowledgement

We extend our sincere thanks to all the participants for their valuable time and contribution to this study.

