## Supplementary material for "Unravelling the Links Between Chronotype, Body Mass Index, and Self-Regulatory Eating Behavior: Preliminary Insights from an Urban Kolkata Study": Table S1

**Table S1: Association between the foods that participants consider tempting in relation to their BMI and employment status**

| Food participants find appealing | BMI | | | | p value |
|---|---|---|---|---|---|
| | **Underweight** | **Normal** | **Overweight** | **Obese** | |
| **Chocolate** | 11 (14.1%) | 24 (30.8%) | 18 (23.1%) | 25 (32.1%) | 0.97 |
| **Chips** | 7 (14%) | 15 (30%) | 9 (18%) | 19 (38%) | 0.6 |
| **Sweets** | 8 (16%) | 13 (26%) | 8 (16%) | 21 (42%) | 0.19 |
| **Bread/ toast** | 7 (33.3%) | 5 (23.8%) | 5 (23.8%) | 4 (19%) | **0.033*** |
| **Pizza** | 8 (14%) | 17 (29.8%) | 9 (15.8%) | 23 (40.4%) | 0.23 |
| **Popcorn** | 6 (20%) | 4 (13.3%) | 9 (30%) | 11 (36.7%) | 0.16 |
| **Frizzy drink** | 9 (19.1%) | 14 (29.8%) | 10 (21.3%) | 14 (29.8%) | 0.56 |
| **Pastries** | 5 (17.9%) | 12 (42.9%) | 3 (10.7%) | 8 (28.6%) | 0.15 |
| **Fried food** | 8 (16.7%) | 14 (29.2%) | 10 (20.8%) | 16 (33.3%) | 0.83 |
| **Cake** | 6 (28.6%) | 7 (33.3%) | 5 (23.8%) | 3 (14.3%) | 0.08 |
| **Biscuit** | 2 (9.5%) | 8 (38.1%) | 4 (19%) | 7 (33.3%) | 0.76 |

| Food participants find appealing | Employment | | | p value |
|---|---|---|---|---|
| | **Students** | **Employed** | **Unemployed** | |
| **Chocolate** | 20 (25.6%) | 27 (34.6%) | 31 (39.7%) | 0.71 |
| **Chips** | 9 (18%) | 21 (42%) | 20 (40%) | 0.36 |
| **Sweets** | 10 (20%) | 15 (30%) | 25 (50%) | 0.41 |
| **Bread/ toast** | 6 (28.6%) | 7 (33.3%) | 8 (38.1%) | 0.81 |
| **Pizza** | 15 (26.3%) | 20 (35.1%) | 22 (38.6%) | 0.70 |
| **Popcorn** | 7 (23.3%) | 10 (33.3%) | 13 (43.3%) | 0.99 |
| **Frizzy drink** | 11 (23.4%) | 20 (42.6%) | 16 (34%) | 0.31 |
| **Pastries** | 8 (28.6%) | 8 (28.6%) | 12 (42.9%) | 0.67 |
| **Fried food** | 10 (20.8%) | 20 (41.7%) | 18 (37.5%) | 0.47 |
| **Cake** | 5 (23.8%) | 5 (23.8%) | 11 (52.4%) | 0.5 |
| **Biscuit** | 6 (28.6%) | 6 (28.6%) | 9 (42.9%) | 0.75 |

Chi-square test used.

*Significant at level 0.05
